# Comparing the COVID-19 pandemic in space and over time in Europe, using numbers of deaths, crude rates and adjusted mortality trend ratios

**DOI:** 10.1101/2020.08.21.20179218

**Authors:** Valentina Gallo, Paolo Chiodini, Dario Bruzzese, Elias Kondilis, Dan Howdon, Jochen Mierau, Raj Bhopal

## Abstract

**Background:** Since COVID-19 was declared a pandemic, attempts have been made to monitor trends over time and to compare countries and regions. Insufficient testing for COVID-19 underestimates the incidence and inflates the case-fatality proportion. Given the age- and sex- distribution of morbidity and mortality from COVID-19, the underlying sex- and age-distribution of a population needs to be accounted for. The aim of this paper is to present a method for monitoring trends of COVID-19 using adjusted mortality trend ratios (AMTR).

**Methods:** Age- and sex-mortality distribution of a reference population composed of the first 14,086 fatalities which occurred before the end of March and were reported in Europe by some countries were used to calculate age- and sex-specific mortality rates per 1,000,000 population. These were applied to each country population to calculate the expected deaths. Adjusted Mortality Trend Ratios (AMTRs) with 95% confidence intervals (C.I.) were calculated for selected European countries from 17/03/2020 to 22/06/2020 by dividing observed cumulative mortality, by expected mortality times the crude mortality of the reference population. These estimated the sex- and age-adjusted mortality for COVID-19 per million population in each country.

**Results:** The cumulative mortality from COVID-19, the crude mortality rates, and the AMTRs were calculated for each country and compared. United Kingdom, Italy, France and Spain registered the highest mortality in Europe. On 22/06/2020 in Europe the total mortality rate from COVID-19 was 352 per 1,000,000 inhabitants; and it was highest in Belgium (850 per 1,000,000 inhabitants) followed by Spain, UK, Italy, Sweden and France. When accounting for the underlying age and sex structure of each country, Belgium remained the single country experiencing the highest AMTR of 929 per million inhabitants on 22/06/2020; however Ireland – which had a CMR in line with the total European population – emerged as having experienced a much more important impact of COVID-19 mortality with an AMTR of 550/million on 22/06/2020, higher than Sweden and Italy.

**Conclusions:** In understanding and managing the pandemic of COVID-19, comparable international data is a priority. Our methods allow a fair comparison of mortality in space and over time. The authors urge the WHO, given the absence of age and sex-specific mortality data for direct standardisation, to adopt this method to estimate the comparative mortality from COVID-19 pandemic worldwide.

**Key message:**

- Comparing trends of the COVID-19 pandemic over time and in space is essential to monitor the disease and compare different local policies
- Using the concept of indirect standardisation we propose a method to effectively compare age- and sex-adjusted mortality rates trends interpretable as deaths for COVID-19 per million inhabitants
- Applying this methods, interesting features of the infection in Europe emerged; e.g. by 22/06/2020 Belgium is the most severely affected country with an AMTR of 929 per million inhabitants, followed by the UK; Ireland and Sweden rank fourth and fifth most affected country in Europe
- The WHO should consider using this method for monitoring the spread of COVID-19, which only requires recording the total number of death from COVID-19 from each country/region

## Introduction

In December 2019, a cluster of pneumonia cases in Wuhan City (China) was identified as having been caused by the SARS-CoV-2 virus, leading to the disease now termed COVID-19. The subsequent global transmission led to the outbreak being classified as a pandemic by the World Health Organisation (WHO) on 11^th^ March 2020^1^. Some of the clinical characteristics of COVID-19 infection (long incubation period, heterogeneity of symptoms, transmission by asymptomatic carriers)^1^–^5^ have contributed to make the estimates of its distribution at population level somewhat challenging. Nonetheless, monitoring the pandemic including international comparisons is of paramount importance to understand the infection dynamics, and to prepare healthcare systems. In addition, this would allow estimating the efficacy of the measures adopted in some countries, but not in others.

When comparing data from different countries it is essential that the metric used relies on data collected in a similar way assuring comparability^6^. There are currently variable policies about testing for COVID-19 leading to undercounts, especially of people showing few symptoms. In countries where there is no community testing policy, only those individuals reaching the hospital are tested in order to protect the healthcare workers and the other patients. This leads to selection bias where the most severe cases only are tested, underestimating incidence and inflating case fatality ratios. Without comprehensive community testing the comparison of incidence or any other metric using the number of “confirmed cases”^7,8^ is inaccurate. Conversely, mortality should suffer from less variability given that it is independent from testing policy, and that it should be recorded fairly consistently across countries given the uniqueness of the clinical picture of the people severely affected by COVID-19.

Given the particular age- and sex- distribution of morbidity and mortality from COVID-19^4,9^, the underlying sex- and age-distribution of a population is of great importance in determining the number of expected cases. Therefore, not adjusting for age undermines meaningful comparison between populations, especially when the age structures differs markedly, such as when comparing lower- with higher-income countries. To date, due to the sudden onset and speed of the pandemic, it has been difficult to compare data coming from different countries partly because basic epidemiological principles (e.g. adjustment for age and sex) have not been applied consistently, the main emphasis having been on the number of cases^6,10^.

Standardised Mortality Trend Ratios (SMTRs) were used to describe the COVID-19 related mortality trends over time across Italian regions by some of us^11^. The aim of this paper is to expand on that approach and to present a method for monitoring trends of burden of COVID-19 based on the concept of indirect standardisation^12^. The method is illustrated by an application to recent data coming from European countries.

## Methods

Some considerations on case definition of COVID-19 and related mortality are reported in Panel 1. The main metrics used in epidemiology to monitor disease occurrence are summarised in Panel 2.

**PANEL 1:**
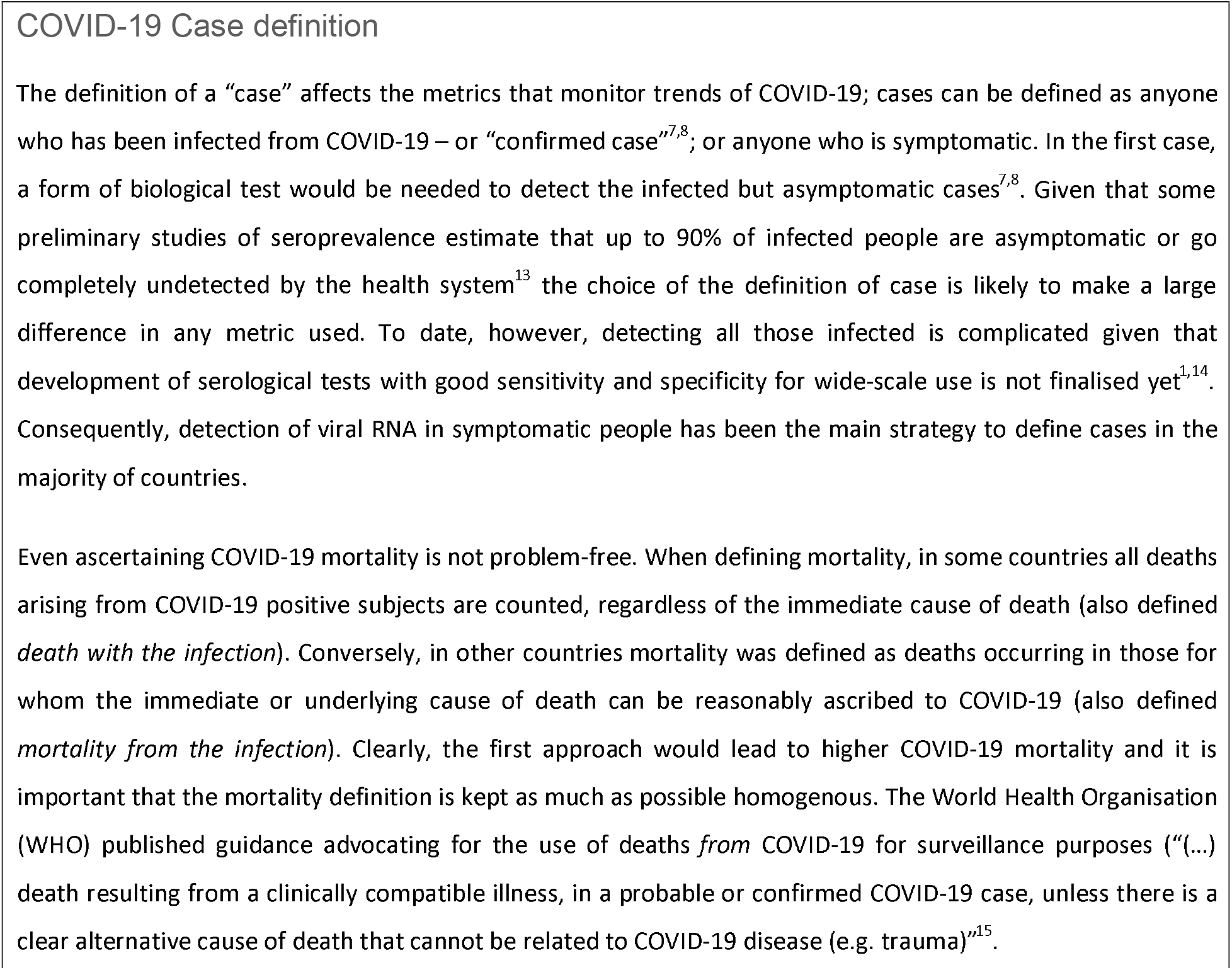
DEFINITION OF COVID-19 CASES AND MORTALITY

**PANEL 2:**
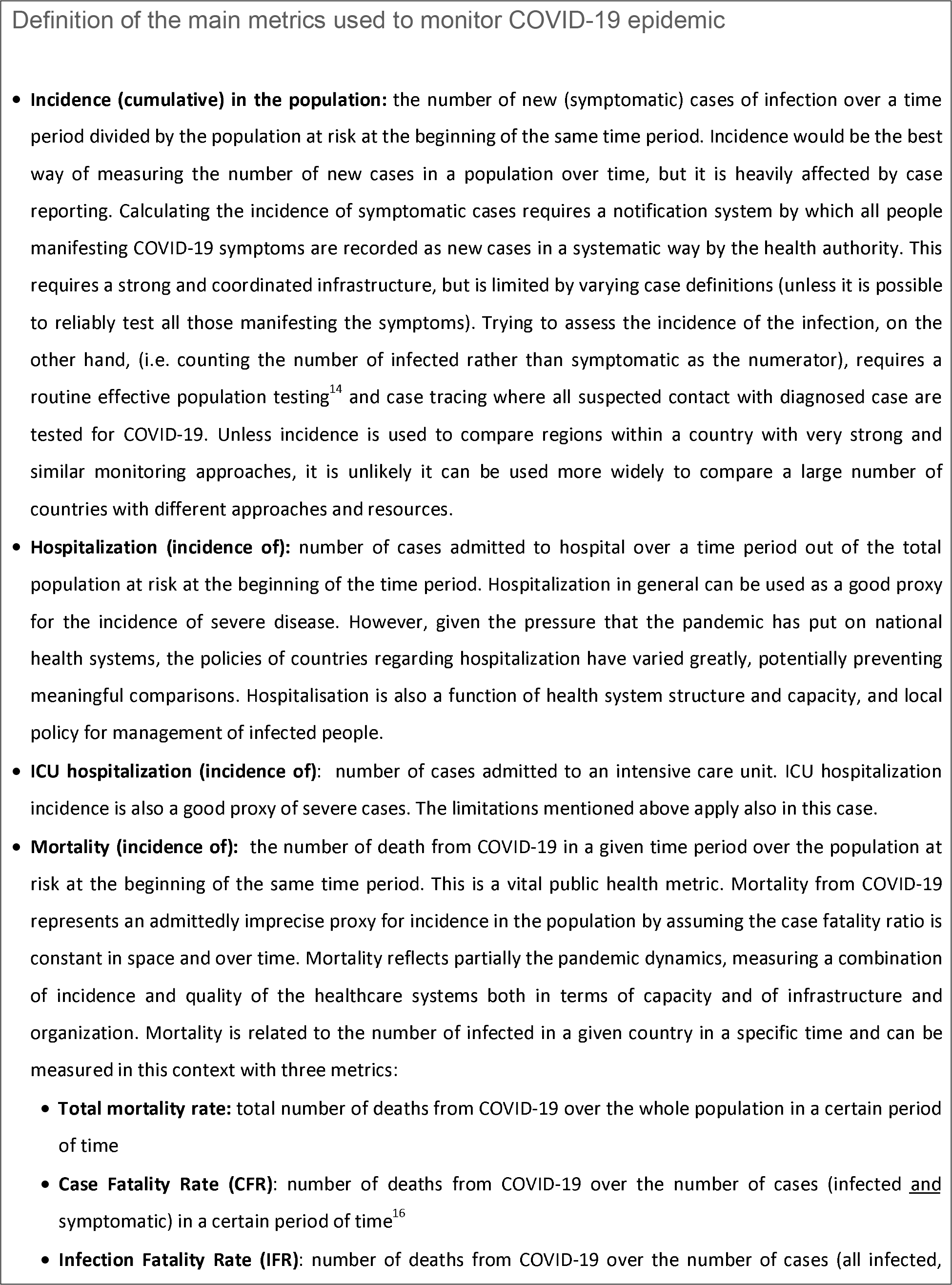
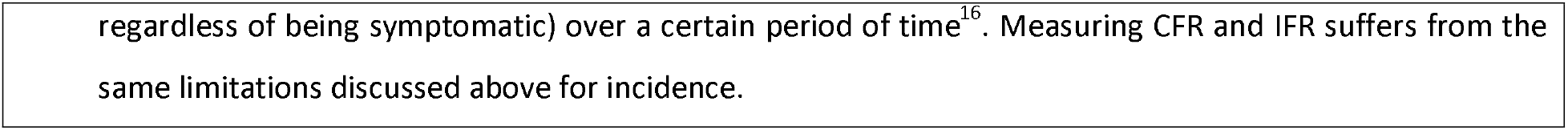
DEFINITION OF THE MAIN METRICS USED TO MONITOR COVID-19

## Data collection and statistical analysis

Daily total number of deaths from COVID-19 from selected European countries from 17/03/2020 to 22/06/2020 were extracted from the European Centre for Disease Prevention and Control (ECDC) website^17^, these were summed up to display cumulative number of deaths per day per country. Crude mortality rate per 1,000,000 was calculated daily in each country by dividing the number of cumulative deaths by the total country population in 2018 extracted from the Organisation for the Economic co-operation and Development (OECD)^18^.

A reference population and a reference period was conveniently defined as the population of the European countries for which the age- and sex-distribution of deaths from COVID-19 by the end of March 2020 was available. Age- and sex distribution of a total of 14,086 COVID-19-related fatalities which occurred during the reference period (649 from the United Kingdom up to 27/03/2020^19^, 4,993 from Italy released on 23/03/2020^20^, 821 from Belgium^17^, 3,459 from France^21^, 581 from Germany^17^, 187 from Portugal^17^, and 3,396 from Spain^17^, all up to 31/03/2020) were used to calculate age- and sex-specific mortality rates per 1,000,000 population. These rates were applied to the entirety of European countries 2018 population in order to estimate the number of expected cases in each country, which can be interpreted as the number each country would have had if they experienced the rates of the reference population by the end of the reference period (end of march 2020)^12^.

For each day, the number of cumulative observed deaths in each country was divided by the number of expected deaths by end of the reference period, and multiplied by 100 to calculate the Standardised Mortality Trend Ratio (SMTR) in the i-th day, as done previously^11^, with the following formula:

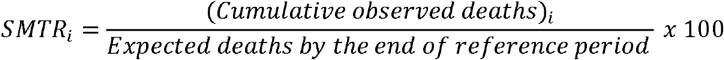

95% confidence intervals of SMTR were calculated assuming a Poisson distribution. Subsequently, each SMTR was multiplied by the crude death rate calculated in the reference period multiplied by 10,000 in order to obtain the Adjusted Mortality Trend Ratio (AMTR) per million inhabitants in the i-th day, applying the following formula, as described in^12^:

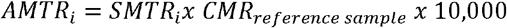

The AMTR can be interpreted as the age- and sex-adjusted number of deaths per million inhabitants due to COVID-19 if the population had experienced the same mortality rate as the reference population in the reference period.

## Results

The daily number of deaths from COVID-19 reported in each country from 17/03/2020 to 22/06/2020 are plotted in in Supplementary Figure 1. While in some countries such as Italy the curve was relatively smooth, in others such as France and Belgium an alternate pattern is observable, probably reflecting some differential timing in reporting of deaths across regions. On 04/04/2020, France reported the overall highest daily mortality, with 2004 deaths from COVID-19 in one day.

Daily cumulative number of deaths was calculated in all European countries which were then ranked as in Table 1. Cumulative deaths were also plotted against time in Figure 1. Italy was the first country registering more than 20,000 deaths in mid-April, and totalled 34,643 deaths by the end of the observation period (22/06/2020). A similar pattern was observed in Spain and France, with only about a week’s delay, which however levelled off reaching a total of 29,927 and 30,037 deaths by 22/06/2020, respectively. Conversely, in the UK, despite the 20,000 deaths threshold being reached about 10 days after Italy, the death toll increase was steady exceeding other countries by 14/05/2020 and registering the highest toll in Europe of 40,585 by the end of the observation period. Belgium, Germany, Netherlands and Sweden registered increasing number of deaths with similar trends, but never reaching the 10,000 deaths threshold.

**Table 1:**
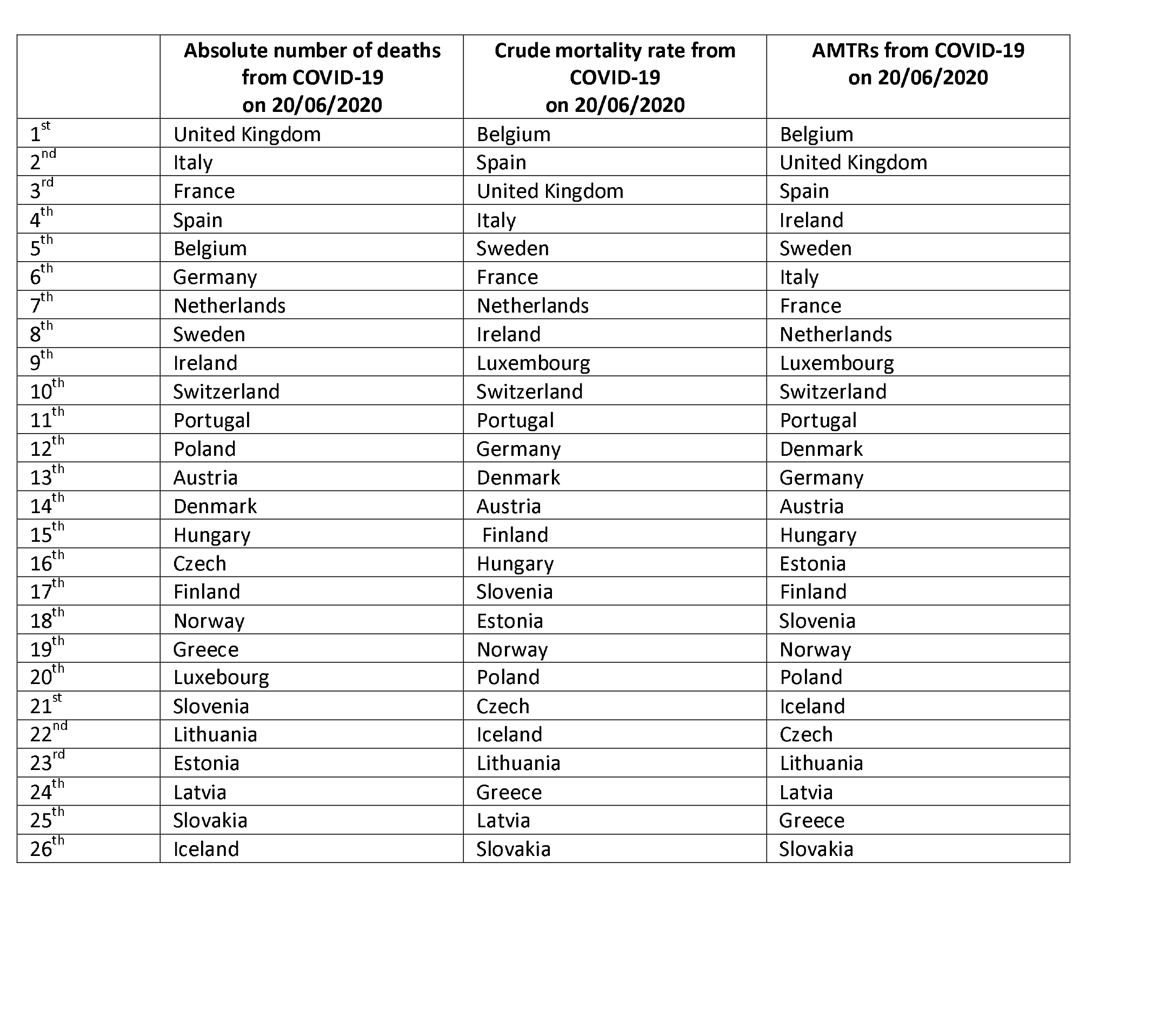
RANKING OF THE INCLUDED EUROPEAN COUNTRIES FOR ABSOLUTE NUMBER OF DEATH FROM COVID-19, CRUDE

|  | **Absolute number of deaths**<br>**from COVID-19**<br>**on 20/06/2020** | **Crude mortality rate from**<br>**COVID-19**<br>**on 20/06/2020** | **AMTRs from COVID-19**<br>**on 20/06/2020** |
| --- | --- | --- | --- |
| 1<sup>st</sup> | United Kingdom | Belgium | Belgium |
| 2<sup>nd</sup> | Italy | Spain | United Kingdom |
| 3<sup>rd</sup> | France | United Kingdom | Spain |
| 4<sup>th</sup> | Spain | Italy | Ireland |
| 5<sup>th</sup> | Belgium | Sweden | Sweden |
| 6<sup>th</sup> | Germany | France | Italy |
| 7<sup>th</sup> | Netherlands | Netherlands | France |
| 8<sup>th</sup> | Sweden | Ireland | Netherlands |
| 9<sup>th</sup> | Ireland | Luxembourg | Luxembourg |
| 10<sup>th</sup> | Switzerland | Switzerland | Switzerland |
| 11<sup>th</sup> | Portugal | Portugal | Portugal |
| 12<sup>th</sup> | Poland | Germany | Denmark |
| 13<sup>th</sup> | Austria | Denmark | Germany |
| 14<sup>th</sup> | Denmark | Austria | Austria |
| 15<sup>th</sup> | Hungary | Finland | Hungary |
| 16<sup>th</sup> | Czech | Hungary | Estonia |
| 17<sup>th</sup> | Finland | Slovenia | Finland |
| 18<sup>th</sup> | Norway | Estonia | Slovenia |
| 19<sup>th</sup> | Greece | Norway | Norway |
| 20<sup>th</sup> | Luxembourg | Poland | Poland |
| 21<sup>st</sup> | Slovenia | Czech | Iceland |
| 22<sup>nd</sup> | Lithuania | Iceland | Czech |
| 23<sup>rd</sup> | Estonia | Lithuania | Lithuania |
| 24<sup>th</sup> | Latvia | Greece | Latvia |
| 25<sup>th</sup> | Slovakia | Latvia | Greece |
| 26<sup>th</sup> | Iceland | Slovakia | Slovakia |

**FIGURE 1:**
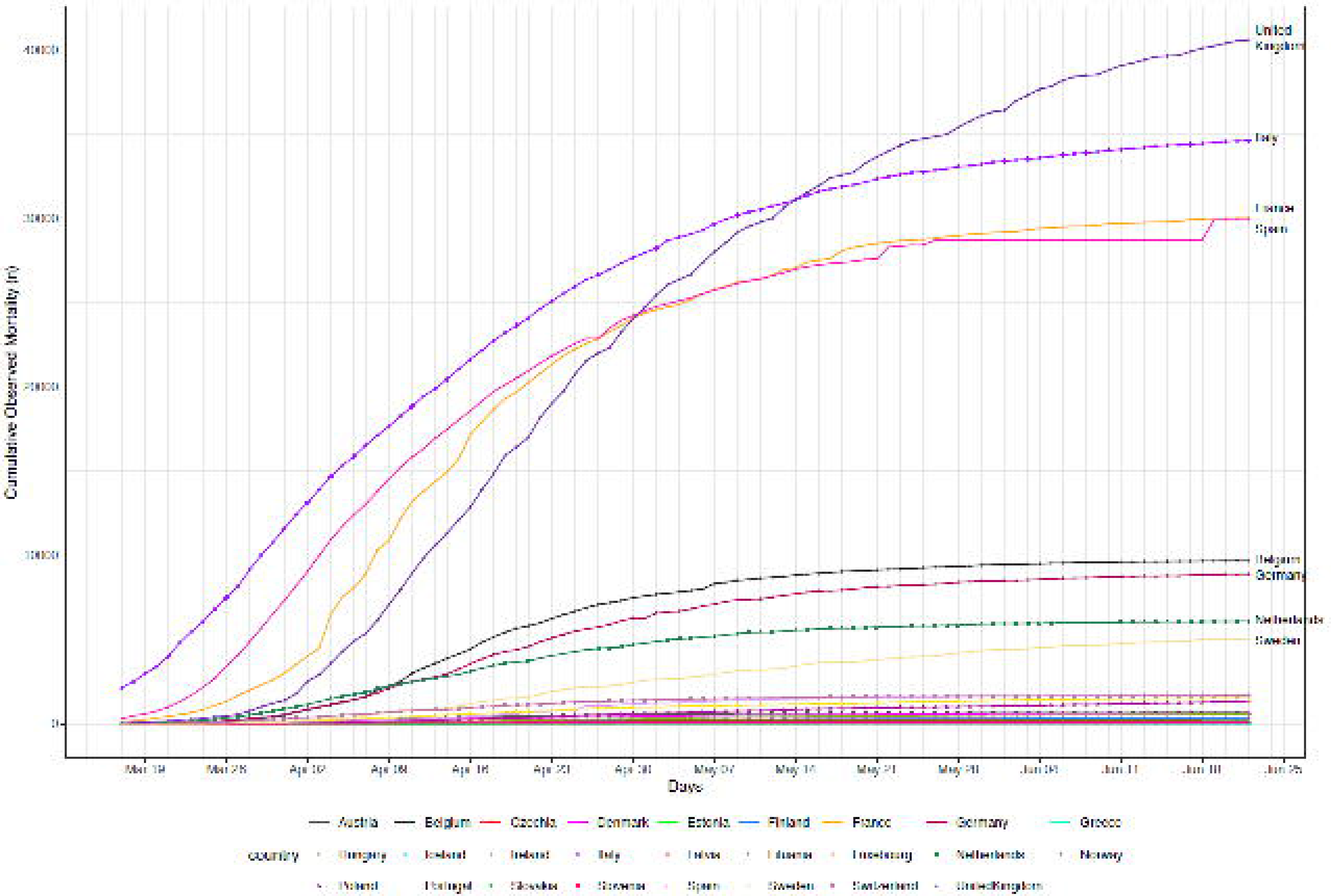
DAILY CUMULATIVE MORTALITY FROM COVID-19 IN SELECTED EUROPEAN COUNTRIES FROM 17/03/2020 TO 22/06/2020

Crude mortality rates were plotted as shown in Figure 2; the resulting ranking of countries by the end of the study period is in Table 1. On 22/06/2020 in Europe the total mortality rate from COVID-19 was 352 per 1,000,000 inhabitants; this was highest in Belgium (850 per 1,000,000 inhabitants on the same date) and lowest in Slovakia (5 deaths per 1,000,000 inhabitants). Once accounting for the total country population, in Belgium there was a sharp increase in crude mortality around the second and third weeks of April, with the highest crude mortality on the 17/04/2020. Spain experienced higher mortality than Italy from the first week of April maintaining higher rates afterwards and reaching a mortality rate of 635 per 1,000,000 inhabitants by 22/06/2020 compared to 573 per 1,000,000 in Italy on the same day. Crude mortality in France was below the Spanish rates, reaching an overall crude mortality rate of 448 by 22/06/2020. The steep increase in mortality in the UK is reflected in the fact that it reached a crude mortality rate of 611 per 1,000,000 inhabitants by the end of observation period. A similar steep pattern in crude mortality rate increase was spotted in Sweden which overtook France by the end of observation period with a crude mortality rate of 497 per 1,000,000 inhabitants. Interestingly, once accounting for the population size, the Netherlands, Ireland, and Luxembourg experienced crude mortality rates aligned with the European average.

**FIGURE 2:**
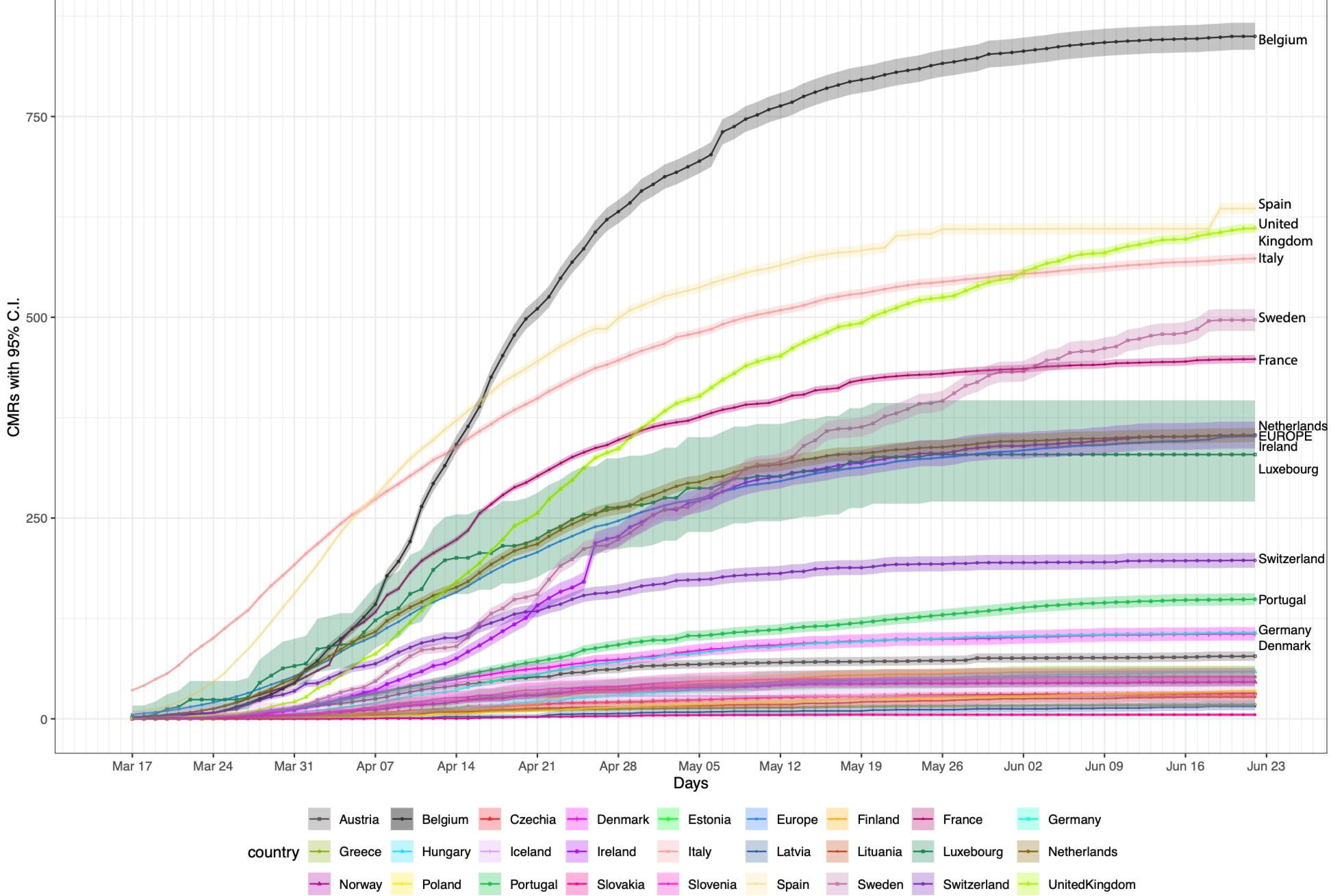
CRUDE MORTALITY RATES (CMRs) FROM COVID-19 PER 1,000,000 INHABITANTS IN EUROPEAN COUNTRIES FROM 17/03/2020 TO 22/06/2020

AMTRs of each country are plotted in Figure 3, and countries were ranked accordingly in Table 1. When accounting for the underlying age and sex structure of each country, the overall trends of the crude mortality ratios were replicated with some notable differences. While Belgium remained the single country experiencing the highest adjusted mortality rates (AMTR on 22/06/2020: 929 per million inhabitants), the United Kingdom was the second ranking European country per age- and sex-adjusted COVID-19 mortality with a AMTR of 698/million on 22/06/2020. Notably, Ireland which had a CMR in line with the total European population (Figure 2), once the underlying sex- and age-structure of the population is accounted for showed a much more important impact of COVID-19 mortality with an AMTR of 550/million on 22/06/2020, higher than Sweden and Italy.

**FIGURE 3:**
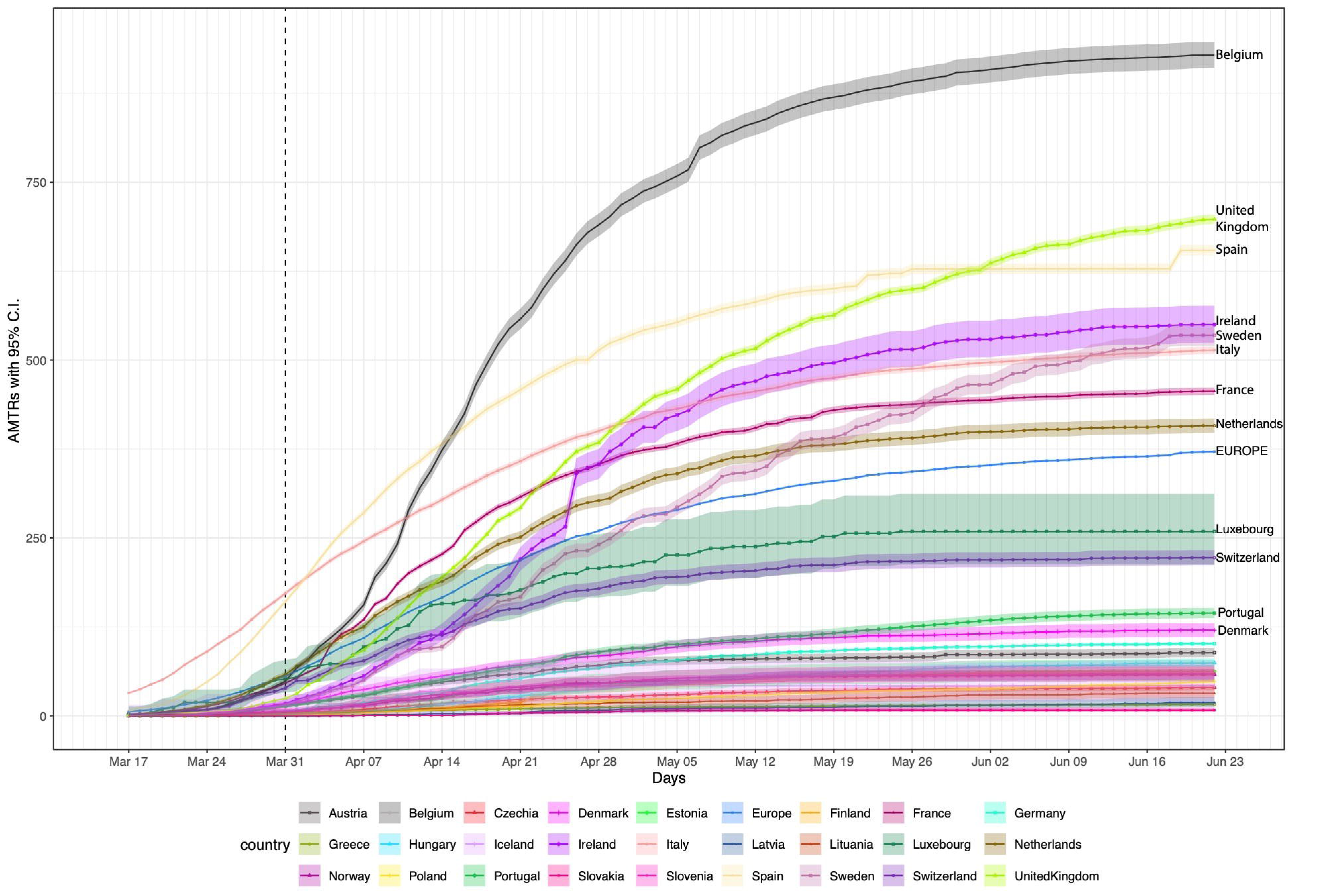
ADJUSTED MORTALITY TREND RATES (AMTRs) PER 1,000,000 INHABITANTS DUE TO MORTALITY FROM COVID-19 IN EUROPEAN COUNTRIES FROM 17/03/2020 TO 22/06/2020

## Discussion

Most reports on COVID-19 epidemiological data to date have not taken the underlying population structures into account, prompting the need for the development of a reliable method for monitoring the COVID-19 pandemic over time and in space while accounting for age-structures of the populations. Using this method of calculating AMTRs, starting from the age- and sex-distribution of mortality among a reference population during a reference period allowed a comparison of trend both in space and over time discovering features of the pandemic otherwise not easily detectable. For example, that United Kingdom experienced the second highest mortality in Europe by the end of June, and that Ireland was the fourth ranking county just above Sweden.

In countries where the social distancing measures have not been taken (i.e. Sweden) or have been substantially delayed (i.e. UK) the curve of AMTRs shows a much steeper shape compared to other European countries which have enforced stricter rules.

## Strengths and limitations

The main strength of this approach for monitoring COVID-19 pandemic is the minimal data requirement to be modelled. By only obtaining periodic COVID-19 total mortality from countries or regions, AMTRs can be calculated allowing a sound comparison of trends in space and over time while accounting for the underlying sex- and age-structure of the population. This allowed comparison between countries with different demographic structures for COVID-19 mortality and other diseases whose mortality is so strongly associated with age.

The main potential limitations of this approach refers to the definition of mortality for COVID-19. Discrepancies in defining deaths from COVID-19 at country/region level would affect the number of deaths reported and therefore the estimated AMTRs. If all authorities would follow the strict guidance provided by the WHO^15^, discrepancies will be minimised reinforcing the reliability of the present method. For example, some scientists have shared concerns on how mortality from COVID-19 is being ascertained in Belgium where people dying in care homes have been classified as dying from COVID-19 based on indirect evidence (the presence of infection in the care home and the reporting of compatible symptoms)^22^. In addition, by calculating the cumulative mortality, any error in death reporting would be carried on in the analysis. Finally, it is important to bear in mind that mortality does not solely reflects the spread of disease: mortality from COVID-19 is function of disease incidence, severity, and quality of healthcare systems to cope with infected and diseased people, therefore caution needs to be exerted when interpreting the AMTRs. Moreover this method assumes that mortality remains constant over time in the given populations. However, it is likely that the age-specific mortality rates calculated during the first phases of the infection reflect an increased mortality of a more vulnerable population exposed to comorbidities. In future similar studies, availability of age- and sex-specific mortality data on a continuing basis, at least from a selected sources, would be useful in order to improve the method by using specific data from across the time course.

## Contextualisation of the results

Reliable comparisons of impact of COVID-19 at regional or national level are essential for the monitoring of the spread of diseases in space and over time, and are instrumental for the retrospective analysis of different non-pharmacological policies currently enforced in many affected countries^23,24^. Nonetheless these are often based on confirmed cases, which suffer greatly from local testing policies^1^, and the intrinsic properties of the serological test.

For example, in a paper comparing confirmed cases in Nigeria and Italy^25^, the difference was striking, but it is unclear how much of it was due to differences in testing policies, and to the deeply different underlying age structure of the populations in the two countries. This is also very relevant when comparing management policies and emergency preparedness: in a study comparing China, Australia, and United States^26^, confirmed cases were used alongside crude mortality, overlooking the importance of the underlying difference in age structure of the populations.

## Conclusions and future directions

In a context of rapid production of scientific evidence aimed at contributing to the understanding and management of a pandemic of devastating proportion such as the COVID-19 one, the release of solid, comparable data should be a high priority. The method proposed in this paper allows comparisons of mortality in space and over time. The authors urge the WHO to adopt this method to estimate the burden of COVID-19 pandemic worldwide given the lack of international data by age and sex that can permit direct standardised rates.

## Data Availability

All data used in this paper are available publicly

---

**SUPPLEMENTARY FIGURE 1:**
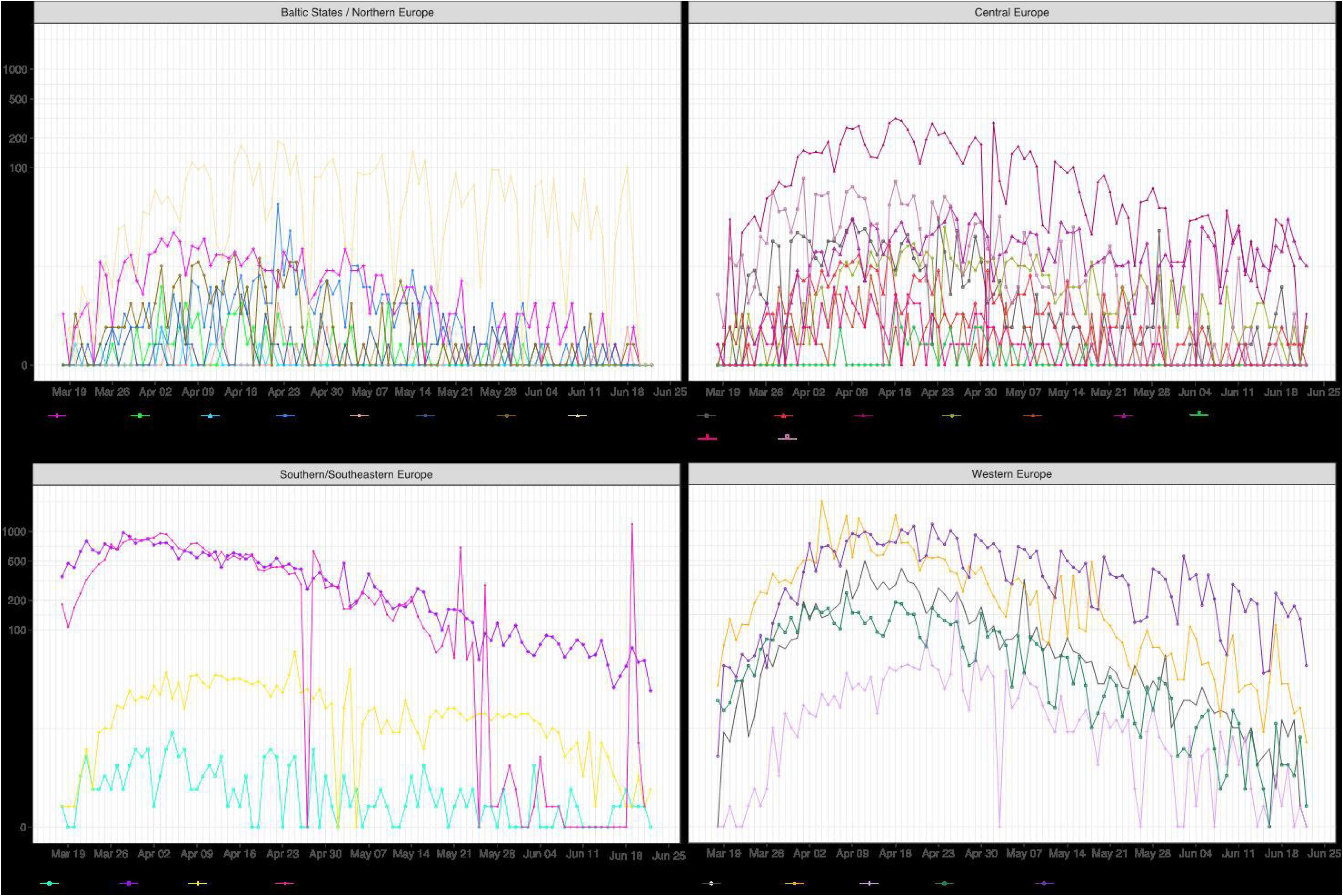
DAILY CUMULATIVE MORTALITY FROM COVID-19 IN SELECTED EUROPEAN COUNTRIES FROM 17/03/2020 TO 22/06/2020

## Notes

### Competing Interest Statement

The authors have declared no competing interest.

### Funding Statement

No specific funding was used to produce this paper

### Author Declarations

No ethics required

